# Onchocerciasis and Non-Communicable Diseases in the Bafut Health District, Cameroon: Knowledge, Attitudes, and Practices Towards Community-Directed Treatment with Ivermectin

**DOI:** 10.1101/2025.09.29.25336865

**Authors:** Irene U. Ajonina-Ekoti, Promise A. Aghaeze, Ebanga Joan, Theophilus A. Ekoti, Tiburce Gangue, Beri A. Gariba, Moses A. Mbanwi, Adolph A. Fozao, Carine K. Nfor, Mbunkah D. Achukwi, Marcelus U. Ajonina

**Affiliations:** Department of Microbiology and Parasitology, The University of Bamenda, Bambili, Cameroon; Department of Educational Psychology, The University of Bamenda, Bambili, Cameroon; Department of Zoology, The University of Bamenda, Bambili, Cameroon; Northwest Regional Delegation of Public Health, Cameroon; District Health Service, Bafut Cameroon; Graduate School of Health and Biomedical Sciences, St Louis University Institute, Douala, Cameroon; TOZARD Laboratory, Bambili, Cameroon; Department of Public Health, The University of Bamenda, Bambili, Cameroon; School of Health Sciences, Charisma University, Billings, Montana, United States of America

**Keywords:** Onchocerciasis, Ivermectin, Community-directed treatment, Knowledge, Attitude, Practices, Non-communicable diseases, Cameroon

## Abstract

**Background:** Onchocerciasis remains a public health problem in Cameroon despite years of community-directed treatment with ivermectin (CDTI). Its persistence increasingly overlaps with the rising burden of non-communicable diseases (NCDs). Community knowledge, attitudes, and practices (KAP) toward onchocerciasis and CDTI, as well as the coexistence of chronic NCDs, may influence the success of elimination programs.

**Methodology:** A community-based cross-sectional study was conducted between April and June 2022 in Bafut Health District, Northwest Cameroon. Using a structured interviewer-administered questionnaire, information was collected on sociodemographic characteristics, knowledge, attitudes, practices related to onchocerciasis and CDTI, and symptoms and comorbidities with NCDs. Descriptive statistics were used to summarize responses, while the association between quantitative variables was determined using the Chi-square test and logistic regression analysis. Statistical significance was set at p<0.05.

**Results:** Of the 250 respondents (mean age 40.9 ± 13.6 years), 96.8% had heard of onchocerciasis, though only 46.0% correctly identified the filarial worm as the cause. Good knowledge, positive attitude, and good practices of onchocerciasis and CDTI were observed in 78.0%, 61.3% and 90.4% of the respondents, respectively. Symptoms of onchocerciasis were reported by 188 participants (75.2%), most commonly itching (50.4%). Of these, 46 (24.5%) also reported an NCD diagnosis, while 84 (44.7%) reported a family history of NCDs. Arthritis (32.4%) was the most common self-reported NCD, and hypertension (14.0%) was the most frequent family history.

**Conclusions:** This study revealed high levels of knowledge and good practices regarding onchocerciasis and CDTI; however, knowledge gaps and concerns about side effects continue to hinder ivermectin uptake. The coexistence of onchocerciasis with chronic NCDs highlights the need for integrated disease management and reinforced health education to support elimination goals.

**Author Summary:** Onchocerciasis, or river blindness, remains a significant public health concern in Cameroon despite decades of community-directed treatment with ivermectin (CDTI). Although CDTI has reduced transmission, the disease continues to affect communities where non-communicable diseases (NCDs) such as arthritis, hypertension, diabetes, stroke, and epilepsy are also on the rise. The coexistence of onchocerciasis and chronic conditions may complicate treatment decisions, affect adherence, and undermine elimination goals. This study was conducted among 250 residents of Bafut Health District, where onchocerciasis is endemic. Using a structured questionnaire, knowledge, attitudes, and practices (KAP) toward onchocerciasis and CDTI were assessed while documenting the occurrence of NCDs. Most respondents demonstrated good knowledge of the disease and reported taking ivermectin. Over 75.2% of participants reported symptoms suggestive of onchocerciasis, and nearly one in four of these also had an NCD diagnosis, while close to half reported a family history of NCDs. Arthritis was the most common condition reported, and hypertension was the most frequent family history. The findings highlight the need for integrated approaches that link onchocerciasis elimination with NCD prevention and management, supported by strengthened community health education.

## Introduction

Onchocerciasis, or river blindness, is a major neglected tropical disease (NTD) that continues to impose a substantial public health and socioeconomic burden. Globally, an estimated 21 million individuals are infected, with more than 99% of cases occurring in sub-Saharan Africa [1]. The disease is caused by the filarial parasite Onchocerca volvulus, transmitted to humans through the bite of infected female blackflies (Simulium spp.), which breed along fast-flowing rivers and streams [2]. During a blood meal, the fly introduces infective third-stage larvae into the human host, where they develop into adult worms that reside in subcutaneous tissues. Female worms can release thousands of microfilariae daily, which are responsible for the dermatological and ocular manifestations of the disease [3]. According to the Global Burden of Disease Study, as of 2017, approximately 20.9 million people were infected, of whom 14.6 million had cutaneous disease and 1.15 million were visually impaired or blind [4].

Cameroon remains one of the most endemic countries, with onchocerciasis present in all ten regions. An estimated six million individuals are infected, and more than half of the population resides in high-risk rural areas along rivers where blackflies are abundant [5,6]. Clinical manifestations include severe itching, skin depigmentation and disfigurement, ocular lesions, and progressive loss of vision. The condition is the second leading cause of infectious blindness worldwide after trachoma. Beyond physical suffering, stigmatization and social exclusion are common, particularly among women and young people, who may face reduced marriage prospects and diminished social participation [7].

Despite decades of control efforts, including vector control and large-scale mass drug administration (MDA) with ivermectin through the community-directed treatment with ivermectin (CDTI) strategy, onchocerciasis persists in several endemic foci. Projections from epidemiological models suggest that elimination by 2030 may not be achievable if MDA remains the sole intervention[8,9]. Several challenges hinder elimination goals: low adherence to ivermectin, treatment refusals due to adverse effects, limited program reach in conflict-affected areas, and interruptions caused by sociopolitical instability and environmental barriers [10,11]. Misconceptions and knowledge gaps about the cause and transmission of onchocerciasis continue to weaken community engagement in control efforts [10,11].

Coinfections and comorbidities also complicate elimination efforts. In particular, ivermectin can trigger severe adverse events in individuals co-infected with Loa loa [12,13]. Additionally, the rising burden of non-communicable diseases (NCDs), including diabetes, hypertension, arthritis, epilepsy, stroke, and cancers, presents new challenges for endemic communities [4,14]. NCDs share complications such as vision loss with onchocerciasis, and anecdotal reports suggest that people with conditions like arthritis often avoid ivermectin due to perceived or real side effects. Recent studies have also demonstrated associations between *O. volvulus* infection and epilepsy in Cameroon and other sub-Saharan countries [15–18]. With NCDs now responsible for more than 70% of global mortality, disproportionately affecting low-and middle-income countries, the intersection between NCDs and onchocerciasis warrants closer attention [4]. In Cameroon, where NCDs account for more than 30% of deaths, community health workers report that up to 40% of ivermectin recipients experience adverse effects, further undermining adherence.

Community perceptions regarding onchocerciasis and its management remain critical for the success of elimination programs. Understanding how endemic populations perceive the disease, its treatment, and its overlap with NCDs is essential for designing sustainable, context-specific strategies. However, little research has systematically explored these issues, particularly in the Bafut Health District. Despite decades of mass drug administration, the overlap of onchocerciasis with rising NCDs in endemic communities remains poorly characterized. This study, therefore, aims to assess community knowledge, attitudes, and practices regarding onchocerciasis, while also examining its comorbidities with NCDs in Bafut, Cameroon.

## Methods

### Ethical considerations

The University of Bamenda Institutional Review Board (No.2022/078H/UBa/IRB), the Northwest Regional Delegation of Public Health and the District Medical Officer (DMO) of Bafut Health District approved the study protocol. Written informed consent was obtained from all respondents, either by signature or thumbprint, after a detailed explanation of the study’s nature and objectives. Participation was entirely voluntary. Data collected was treated with strict confidentiality and stored on password-protected computers. All participants were handled per the principles of the Declaration of Helsinki on the ethical use of humans in biomedical research.

### Study setting and population

This was a community-based cross-sectional study conducted at the Bafut Health District (BHD) from April to June 2022. The BHD is located in the Mezam Division (6°10′0.000″N, 10°6′0.000″E), Northwest Region, about 20km from Bamenda town along a 10-kilometre stretch of the “Ring Road” that trails along a ridge above the Menchum Valley[19]. The BHD is one of the twenty-one health districts in the northwest region[20], with fourteen health areas. According to the 2004 Rapid Epidemiological Assessment (REA) of onchocerciasis by the Bafut District Health Service, these health areas are classified as hypo-endemic, meso-endemic, or hyper-endemic [21]. The district has an estimated population of over 70,000 inhabitants, predominantly rural, with farming, fishing, hunting, trading, and small-scale businesses being the major sources of livelihood [22].

Bafut is the most powerful of the traditional kingdoms of the Grassfields, now divided into 26 wards. Administratively, the Bafut Sub-Division is one of the five sub-divisions in the Mezam Division, covering a land area of approximately 340 km² (Fig 1). Geographically, it lies within the western grassfields ecological zone, which extends across the Northwest Region of Cameroon and neighbouring highland savannah areas. The River Mezam and its tributaries drain the sub-division: the central collector and its tributaries form a relatively sparse, rectangular drainage network. The presence of ecological features such as forested landscapes with fast-flowing rivers creates suitable habitats for Onchocerca vectors, contributing significantly to the persistence of onchocerciasis. The area also experiences high rainfall, elevated temperatures, and intense human activity, all of which further support vector proliferation and disease persistence.

**Fig 1.**
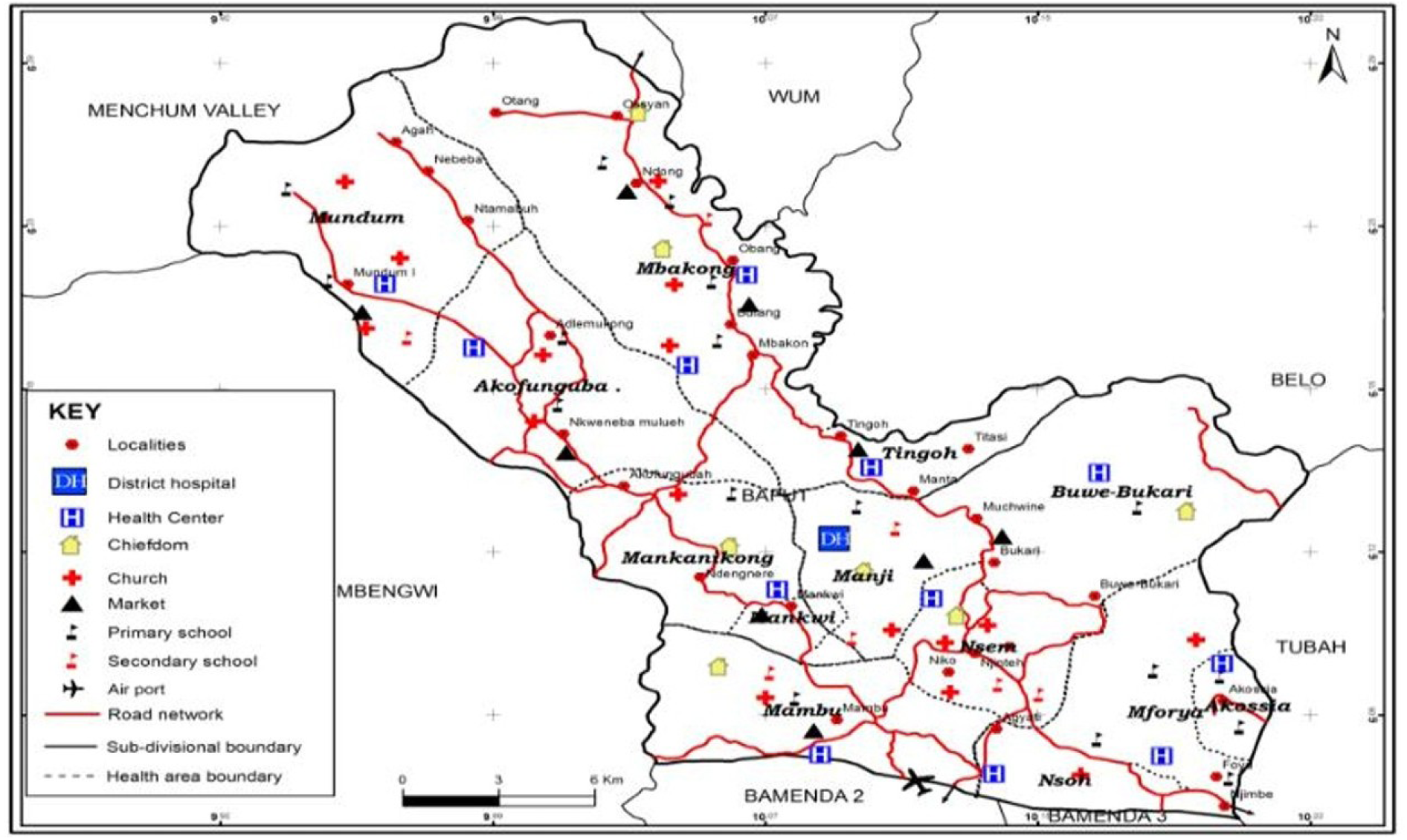
Map of Bafut Health District [19]. The study population comprises adult residents (≥18 years) who have lived in the community for at least two years and were willing to participate by signing an informed consent.

### Sample size and sampling method

The minimum sample size was determined using Lorentz’s formula at a 95% confidence level and 5% margin of error as follows:

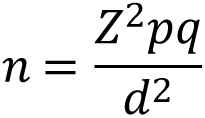

Where,

Where z^2^ = (1.96)2, p = previous knowledge of onchocerciasis and q = 1-p, d^2^ = (0.05)^2^. Domche et al [23] reported 91.6% knowledge level(p) regarding onchocerciasis and black fly nuisance and bio-ecology in a study conducted in the centre region of Cameroon, resulting in a minimum of 160 participants. Households across the health areas of the BHD were randomly selected, and one adult member from each household (≥18 years of age and resident for at least two years) was interviewed. To account for potential non-response, 10% was added to the calculated sample size, giving a minimum requirement of 160 participants.

### Data collection instrument

Data was collected using a pre-tested structured interviewer-administered questionnaire developed from previous KAP studies on onchocerciasis [17,23–25]and adapted to include NCD-related components (supplementary file no. 1) and used to document information on sociodemographic characteristics of participants, Knowledge of onchocerciasis and community-directed treatment with ivermectin (CDTI), attitudes and practices toward onchocerciasis and CDTI, and Symptoms and NCDs. The questionnaire consisted of 31 items. Eight items captured sociodemographic information that included gender, age, education level, marital status, religion, health area (endemicity) and duration of residence in the community.

Nine items assessed knowledge of onchocerciasis and CDTI, including awareness, transmission, symptoms, prevention, ivermectin distribution, and management of side effects. Five items measured attitudes, covering perceptions of disease severity, ivermectin effectiveness, acceptance of annual treatment, and the role of traditional medicine. Four items addressed practices, focusing on ivermectin use, timing of last intake, side effects, and reasons for refusal. The final five items explored symptoms and NCDs, documenting self-reported onchocerciasis symptoms, prior diagnoses of major NCDs, and relevant family history.

### Variables

#### Dependent variables

The outcome variables for this study were knowledge of onchocerciasis and CDTI, attitudes toward the disease and its treatment, preventive practices, and reported onchocerciasis-NCD comorbidity. Knowledge was assessed using eight core questions covering awareness, transmission, symptoms, prevention, and ivermectin distribution. A respondent who had correct responses for at least five of the components was considered to have good knowledge, while those with correct responses to four or fewer were considered to have poor knowledge regarding onchocerciasis and CDTI.

Attitudes were assessed using five items: perception of onchocerciasis as a serious disease, belief in the effectiveness of ivermectin, willingness to encourage family or friends to take ivermectin, comfort with annual ivermectin intake, and reliance on traditional medicine. A response of “Yes” to items on disease seriousness, ivermectin effectiveness, willingness to encourage others, and comfort with annual intake was coded as positive, while “No” or “I don’t know” were coded as negative. For the traditional medicine item, “No” was coded as positive, while “Yes” and “I don’t know” were coded as negative. A respondent with three or more positive responses was classified as having an overall positive attitude; otherwise, the respondent was classified as negative.

Practices were assessed using six items, of which three were used for scoring. These included whether respondents had ever taken ivermectin, the timing of the most recent dose, and willingness to take ivermectin anytime recommended. Ever taking ivermectin, taking it within the past year or a few months, and being willing to take it anytime recommended were coded as right practices, while alternative responses were coded as wrong practices. Experience of side effects and the type of symptoms reported were treated descriptively and not classified as right or wrong. A respondent with two or more right practices was classified as having good practice; otherwise, poor practice. The cut-off points used for classifying good knowledge, positive attitude, and good practices were adapted from similar KAP studies in Cameroon and Nigeria [23,24].

The comorbidity domain captured self-reported symptoms of onchocerciasis (itching, nodules, blurred vision, skin changes) alongside prior diagnosis or family history of selected NCDs, including hypertension, diabetes, stroke, arthritis, and epilepsy.

#### Independent variables

Explanatory variables were limited to the socio-demographic factors including; gender (male, female), age group (<30, 31–50, >50 years), marital status (single, married), level of education (none, primary, secondary, tertiary), occupation (student, farmer, employed, business), religion (Christian, Muslim, Atheist), duration of residence in the area (<2 years, 3–10 years, >10 years), and health area endemicity (hypo-, meso-, hyper-endemic).

### Data analysis

All completed questionnaires were checked for completeness, coded, and entered into Microsoft Excel before being exported to SPSS Statistics version 27.0 (IBM Corp., Armonk, NY, USA) for analysis. Descriptive statistics such as frequencies, percentages, means, and standard deviations were used to summarise socio-demographic characteristics, knowledge, attitudes, practices, and reported symptoms and NCDs.

Associations between categorical variables, including socio-demographic factors and knowledge, attitude and practices regarding onchocerciasis and CDTI, were examined using Pearson’s χ² test. To determine factors associated with good knowledge, positive attitude and good practices towards onchocerciasis and CDTI, logistic regression analysis was performed. Variables with a p-value ≤0.10 in bivariate analysis were entered into the multivariable logistic regression model. Adjusted Odds Ratios (AOR) with their 95% confidence intervals (CI) were calculated to assess the strength of association. Statistical significance was set at p < 0.05.

## Results

A total of 250 participants with a mean age (±SD) of 40.96 ± 13.59 years were enrolled in the study. The majority were female (54.0%), within the 31–50-year age group (50.4%), married (65.2%), had a secondary level of education (64.0%), of the Christian religion (88.0%), with farming as the predominant occupation (49.6%) (Table 1). Most of the participants reported having lived in the area for more than 10 years (69.6%), especially in meso-endemic areas (46.4%).

**Table 1.**
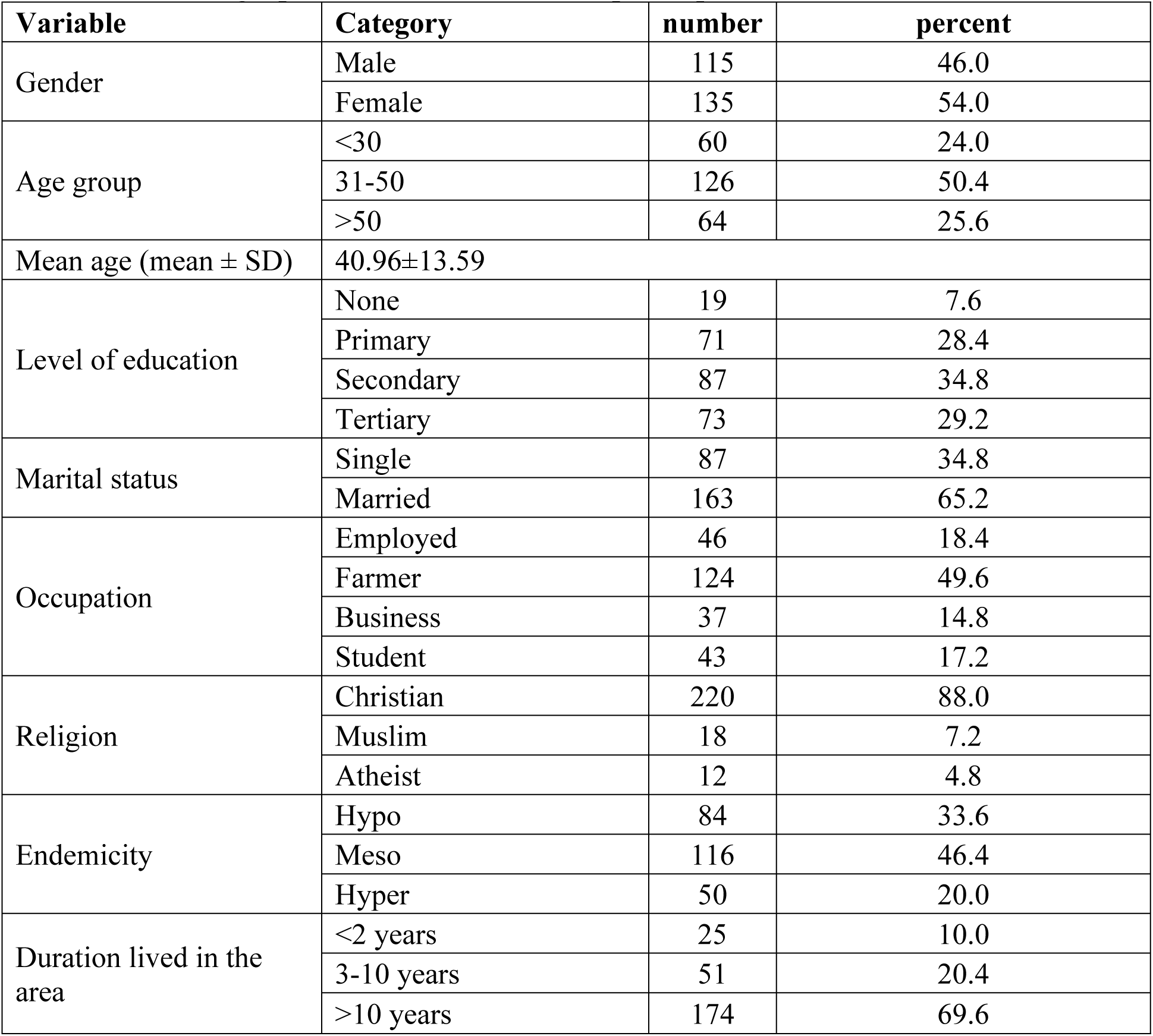
Sociodemographic characteristics of the participants (N=250).

### Knowledge regarding onchocerciasis and CDTI

Almost all participants (96.8%) acknowledged having heard about onchocerciasis (Table 2), however, only 46.0% correctly identified the filarial worm as the cause of the disease, while 35.6% attributed it to blackflies, and 12.4% to mosquitoes; a small proportion linked it to poor hygiene (1.6%). Regarding transmission, the majority (58.8%) knew that blackfly bites are responsible, but 16.0% wrongly attributed mosquito bites. On clinical presentation, itching (66.0%), skin changes (32.0%), and nodules (37.6%) were commonly recognised, though only 8.4% mentioned oedema. When asked about the possibility of person-to-person transmission of onchocerciasis, 38.3% of respondents believed it was possible, whereas 59.2% correctly indicated that it was not, and 2.5% reported being uncertain.

**Table 2.** Knowledge of Onchocerciasis and CDTI (N=250).

| **Indicative questions** | **Response categories** | **n(%)** |
| --- | --- | --- |
| Have you ever heard about onchocerciasis? | Yes | 242(96.8) |
|  | No | 8(3.2) |
| What causes onchocerciasis?* | Filarial worm | 115(46.0) |
|  | Blackfly | 89(35.6) |
|  | Mosquito | 31(12.4) |
|  | Poor personal hygiene | 4(1.6) |
|  | I don't know | 11(4.4) |
| How are people infected with the disease?* | Blackfly bite | 147(58.8) |
|  | Contact with infected persons | 21(8.4) |
|  | Mosquito bite | 40(16.0) |
|  | Filarial worm | 4(1.6) |
|  | I don't know | 38(15.2) |
| What are the signs and symptoms of onchocerciasis? * | itching | 165(66.0) |
|  | edema | 21(8.4) |
|  | skin change | 80(32.0) |
|  | Nodules in any part of the body | 94(37.6) |
|  | I don't know | 23(9.2) |
| Can onchocerciasis be transmitted from person to person? | Yes | 92(38.3) |
|  | No | 148(59.2) |
|  | I don't know | 10(2.5) |
| Is onchocerciasis preventable? | Yes | 214(85.6) |
|  | No | 15(6.0) |
|  | I don't know | 21(8.4) |
| If yes, how can onchocerciasis be prevented? * | Wearing protective clothing | 68(27.5) |
|  | Avoiding bathing in rivers | 47(18.8) |
|  | Drugs (Ivermectin) | 130(50.6) |
|  | Environmental sanitation | 58(23.5) |
|  | Bednets | 38(15.4) |
|  | I don't know | 15(6.0) |
| Are you aware of the annual ivermectin distribution? | Yes | 224(89.6) |
|  | No | 26(10.4) |
| Do you know that the side effects of ivermectin are treated free of charge? | Yes | 186(83.4) |
|  | No | 37(16.4) |

\*Percentages may exceed 100% due to multiple responses.

Awareness of prevention was generally high, with 85.6% of respondents acknowledging that onchocerciasis can be prevented, while 6.0% considered it not preventable and 8.4% were uncertain. The primary source of information on ivermectin distribution was community drug distributors (CDDs) (46.4%), while neighbours accounted for the lowest proportion (2.4%) (Fig 2). Regarding preventive measures, 50.6% cited ivermectin, 27.5% mentioned the use of protective clothing, 18.8% referred to avoiding bathing in rivers, 23.5% indicated environmental sanitation, and 15.4% suggested the use of bed nets. Awareness of annual ivermectin distribution was widespread (89.6%), and a majority (83.4%) were also aware that any side effects of ivermectin are managed free of charge.

**Fig 2.**
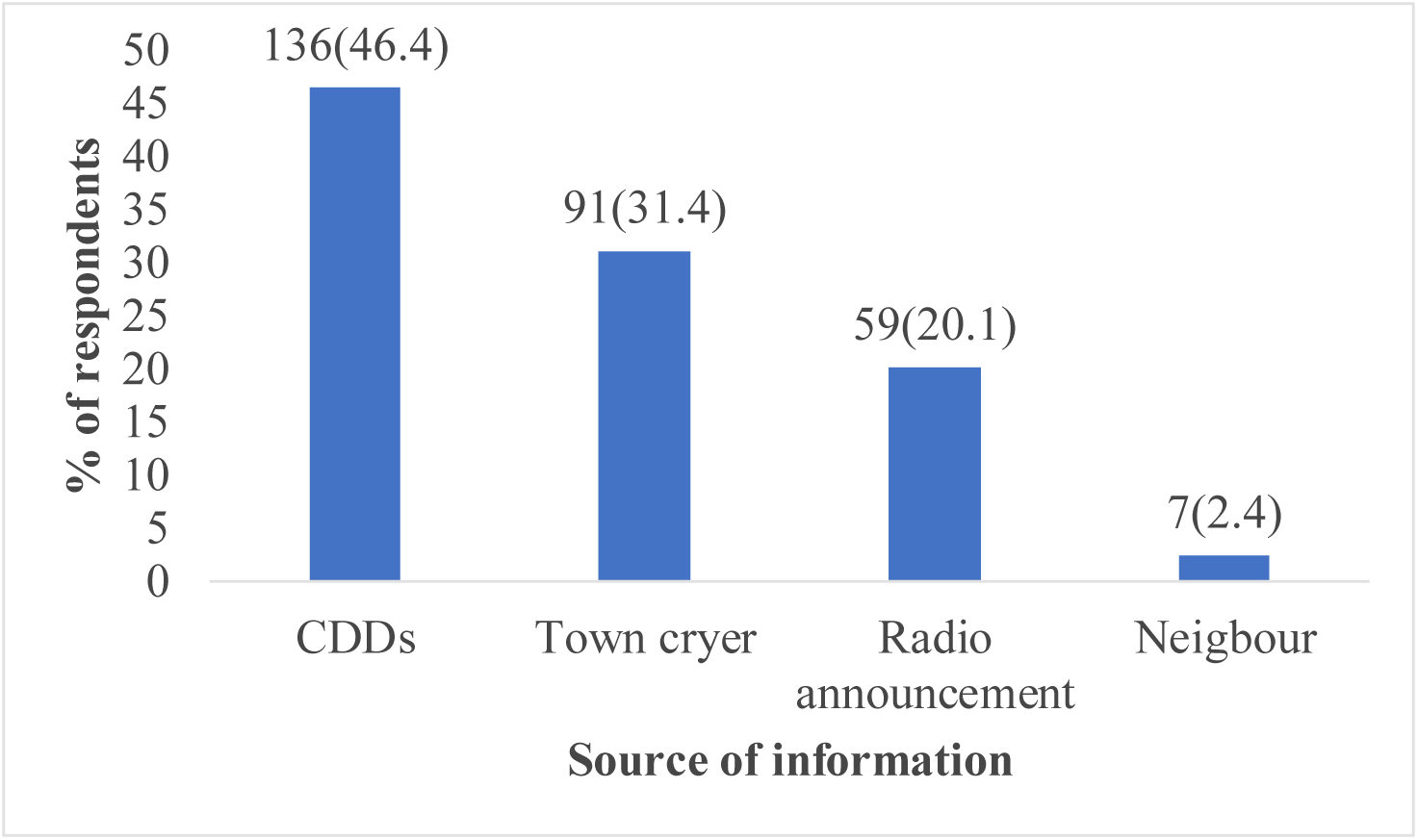
Source of information on ivermectin distribution among respondents.

Overall, the majority, 195(78.0%) of respondents had a good level of knowledge, while 55(22.0%) had a poor level of knowledge on onchocerciasis and CDTI. Analysis revealed that age, education, and endemicity were significantly associated with knowledge of onchocerciasis and CDTI (p<0.05). Multivariate analysis showed that respondents aged over 50 years had the highest level of good knowledge (95.3%) and were six times more likely to demonstrate good knowledge compared to those under 30 years (aOR = 5.62, 95% CI: 1.51–20.88, p = 0.010). In contrast, participants aged 31–50 were less likely to report good knowledge (aOR = 0.42, 95% CI: 0.21– 0.86, p = 0.017) (Table 3). Moreover, our results further showed that educational attainment is also positively associated with knowledge. Participants with tertiary education had the highest knowledge levels (86.3%) and were almost six times more likely to demonstrate good knowledge than those without formal education (aOR = 5.67, 95% CI: 1.85–17.39, p = 0.002). Similarly, respondents with primary education were more than three times as likely to report good knowledge (aOR = 3.66, 95% CI: 1.25–10.72, p = 0.018).

**Table 3.** Association between knowledge of onchocerciasis/CDTI and sociodemographic characteristics.

| Variable | Category | N | Good knowledge<br>[n (%)] | Unadjusted<br>p-value | OR | 95%CI | Adjusted<br>p-value |
| --- | --- | --- | --- | --- | --- | --- | --- |
| Gender | Male | 115 | 90(78.3) | 0.817 | NA |  |  |
|  | Female | 135 | 104(77.0) |  |  |  |  |
| Age group | <30 | 60 | 47(78.3) | <0.001 | 1 |  |  |
|  | 31-50 | 126 | 76(60.3) |  | 0.42 | 0.21 – 0.86 | 0.017* |
|  | >50 | 64 | 61(95.3) |  | 5.62 | 1.51 – 20.88 | 0.010* |
| Level of education | None | 19 | 10(52.6) | 0.011* | 1 |  |  |
|  | Primary | 71 | 57(74.6) |  | 3.66 | 1.25 – 10.72 | 0.018* |
|  | Secondary | 87 | 61(70.1) |  | 2.11 | 0.77 – 5.80 | 0.147 |
|  | Tertiary | 73 | 63(86.3) |  | 5.67 | 1.85 – 17.39 | 0.002 |
| Marital status | Single | 87 | 70(80.5) | 0.428 | NA |  |  |
|  | Married | 163 | 124(76.1) |  |  |  |  |
| Occupation | Student | 43 | 36(83.7) | 0.068 | 1 |  |  |
|  | Employed | 46 | 37(80.4) |  | 0.8 | 0.27–2.37 | 0.686 |
|  | Farmer | 124 | 88(71.0) |  | 0.48 | 0.19–1.23 | 0.127 |
|  | Business | 37 | 33(89.2) |  | 1.61 | 0.47–5.53 | 0.446 |
| Religion | Christian | 220 | 171(77.7) | 0.143 | NA |  |  |
|  | Muslim | 18 | 16(88.9) |  |  |  |  |
|  | Atheist | 12 | 7(58.3) |  |  |  |  |
| Endemicity | Hypo | 84 | 73(86.9) | 0.026* | 3.12 | 1.31 – 7.45 | 0.010* |
|  | Meso | 116 | 87(75.0) |  | 1.41 | 0.68 – 2.92 | 0.353 |
|  | Hyper | 50 | 34(68.0) |  | 1 |  |  |
| Duration lived in the area | <2 years | 25 | 20(80.0) | 0.379 | NA |  |  |
|  | 3-10 years | 51 | 43(84.3) |  |  |  |  |
|  | >10 years | 174 | 131(75.3) |  |  |  |  |

Endemicity level of residence was strongly associated with knowledge, with respondents in hypo-endemic areas being over three times more likely to report good knowledge compared to those in hyper-endemic areas (OR = 3.12, 95% CI: 1.31–7.45, p = 0.010). Those in meso-endemic areas also had higher knowledge (75.0%) relative to hyper-endemic regions, though this difference was not statistically significant (p<0.05).

### Attitudes toward Onchocerciasis and CDTI

Attitude of respondents towards onchocerciasis and CDTI was assessed using five questions (Table 4). Of all the respondents, less than half (39.2%) considered onchocerciasis a severe disease, while 44.0% did not, and 16.8% were uncertain (Table 4). Similarly, 43.0% of respondents perceived ivermectin as effective in preventing onchocerciasis, while 32.2% disagreed and 24.8% were unsure.

**Table 4.** Attitude of the respondent towards onchocerciasis and CDTI (N=250).

| Indicative questions | Response categories | n (%) |
| --- | --- | --- |
| Do you think onchocerciasis is a serious disease in your community? | Yes | 98(39.2) |
|  | No | 110(44.0) |
|  | I don't know | 42(16.8) |
| Do you think Ivermectin is effective in preventing onchocerciasis | Yes | 108(43.0) |
|  | No | 81(32.2) |
|  | I don't know | 61(24.8) |
| Would you encourage family/friends to take ivermectin during CDTI? | Yes | 141(56.4) |
|  | No | 109(43.6) |
| Do you think traditional medicine can cure onchocerciasis? | Yes | 81(32.4) |
|  | No | 109(43.5) |
|  | I don't know | 60(24.1) |
| Are you comfortable taking ivermectin every year? | Yes | 219(88.3) |
|  | No | 31(11.7) |

It was further observed that most respondents (88.3%) reported being comfortable with taking ivermectin annually, and over half (56.4%) indicated they would encourage family or friends to participate in CDTI. However, perceptions of the seriousness of onchocerciasis and the effectiveness of ivermectin were more divided, with fewer than half recognising the disease as severe (39.2%) or affirming ivermectin’s preventive value (43.0%). Notably, nearly one-third (32.4%) believed traditional medicine could cure onchocerciasis, reflecting persistent misconceptions.

It was generally observed that 153 participants (61.3%) demonstrated a positive attitude, while 97 (38.7%) exhibited a negative attitude toward onchocerciasis and CDTI. Positive attitude toward onchocerciasis and CDTI was significantly associated with gender, education, religion, and endemicity (p < 0.05) (Table 5). Female respondents were 1.3 times more likely to demonstrate a positive attitude than males (aOR = 1.35, 95% CI = 1.02–2.45, p = 0.041). Participants with tertiary education were over three times more likely to have a positive attitude than those without formal education (aOR = 3.12, 95% CI = 1.45–6.74, p = 0.003). Respondents of the Muslim faith were also more likely to show positive attitudes than Christians (aOR = 2.45, 95% CI = 1.02–7.88, p = 0.037). In addition, residing in hyperendemic areas was significantly associated with higher odds of positive attitude compared to hypoendemic areas (aOR = 1.92, 95% CI = 1.12–4.35, p = 0.029). Other factors, including age, marital status, occupation, and duration of residence, were not significantly associated with attitude.

**Table 5.** Association between attitude towards onchocerciasis/CDTI and sociodemographic characteristics.

| Variable | Category | N | Positive attitude n (%) | Unadjusted p-value | OR | 95% CI | Adjusted p-value |
| --- | --- | --- | --- | --- | --- | --- | --- |
| Gender | Male | 115 | 82 (71.3) | 0.042* | 1 |  |  |
|  | Female | 135 | 104 (77.0) |  | 1.35 | 1.02–2.45 | 0.041* |
| Age group | <30 | 60 | 44 (73.3) | 0.118 | NA |  |  |
|  | 31–50 | 126 | 91 (72.2) |  |  |  |  |
|  | >50 | 64 | 48 (75.0) |  |  |  |  |
| Education level | None | 19 | 10 (52.6) | <0.001* | 1 |  |  |
|  | Primary | 71 | 48 (67.6) |  | 1.45 | 0.71–3.13 | 0.291 |
|  | Secondary | 87 | 61 (70.1) |  | 1.61 | 0.82–3.52 | 0.184 |
|  | Tertiary | 73 | 63 (86.3) |  | 3.12 | 1.45–6.74 | 0.003* |
| Marital status | Single | 87 | 67 (77.0) | 0.314 | NA |  |  |
|  | Married | 163 | 113 (69.3) |  |  |  |  |
| Occupation | Student | 43 | 31 (72.1) | 0.229 | NA |  |  |
|  | Employed | 46 | 35 (76.1) |  |  |  |  |
|  | Farmer | 124 | 86 (69.4) |  |  |  |  |
|  | Business | 37 | 29 (78.4) |  |  |  |  |
| Religion | Christian | 220 | 159 (72.3) | 0.037* | 1 |  |  |
|  | Muslim | 18 | 16 (88.9) |  | 2.45 | 1.02–7.88 | 0.037* |
|  | Atheist/Other | 12 | 7 (58.3) |  | 0.81 | 0.32–2.15 | 0.665 |
| Endemicity | Hypo | 84 | 61 (72.6) | 0.026* | 1 |  |  |
|  | Meso | 116 | 84 (72.4) |  | 1.08 | 0.65–1.86 | 0.552 |
|  | Hyper | 50 | 42 (84.0) |  | 1.92 | 1.12–4.35 | 0.029* |
| Duration lived in the area | <2 years | 25 | 19 (76.0) | 0.221 | NA |  |  |
|  | 3–10 years | 51 | 39 (76.5) |  |  |  |  |
|  | >10 years | 174 | 126 (72.4) |  |  |  |  |

### Practices related to onchocerciasis prevention

In this study, the majority of respondents (93.2%) reported ever taking ivermectin, while 6.8% had never taken the drug (Table 6). Concerning the timing of uptake, 46.4% had last taken ivermectin during the previous year and 34.8% within a few months, whereas 10.0% had taken it two years prior. Among those who had ever taken the drug, 74.6% reported experiencing some form of side effect after treatment, while 25.4% did not. The most commonly reported side effects were body or eye itching (45.3%), joint pains (25.9%), body swelling (25.3%), and fever (16.5%). Of the 22 respondents who refused to take ivermectin, the main reasons cited were fear of side effects (68.4%), no longer having symptoms of onchocerciasis (18.2%), and a perception that treatment was unnecessary (13.6%). In addition, most respondents (87.6%) indicated they would be willing to take ivermectin anytime it is recommended, while 12.4% expressed unwillingness.

**Table 6.** Practices regarding onchocerciasis prevention.

| Indicative questions | Response categories | N | n (%) |
| --- | --- | --- | --- |
| Have you ever taken Mectizan (Ivermectin)? | Yes | 250 | 233(93.2) |
|  | No |  | 22(6.8) |
| When was the last time you took ivermectin? | A few months ago | 228 | 87(34.8) |
|  | Last year |  | 116(46.4) |
|  | The last two years |  | 25(10.0) |
| Did you experience any side effects after taking ivermectin? | Yes | 228 | 170(74.6) |
|  | No |  | 58(25.4) |
| If yes, which side effects did you experience? | Body/eye itching | 170 | 77(45.3) |
|  | Body swelling |  | 43(25.3) |
|  | Joint pains |  | 44(25.9) |
|  | Fever |  | 28(16.5) |
| If you ever refused ivermectin, what was your reason? | Fear of side effects | 22 | 15(68.4) |
|  | No longer have symptoms of onchocerciasis. |  | 4(18.2) |
|  | Did not see the need |  | 3(13.6) |
| Will you be willing to take ivermectin anytime recommended? | Yes | 250 | 219(87.6) |
|  | No |  | 31(12.4) |

The majority of respondents [226(90.4%)] demonstrated good practices, while 24(9.6%) showed poor practices toward onchocerciasis prevention. Binary logistic regression revealed that age, endemicity, and community residence duration were significantly associated with practice level (Table 7). Respondents aged above 50 years were more likely to demonstrate good practices than those younger than 30 (aOR = 3.13, 95% CI: 1.04–9.39, p = 0.041). Living in hypoendemic areas was associated with better practices than residing in hyperendemic areas (aOR = 0.26, 95% CI: 0.07–0.91, p = 0.034). In addition, individuals who had lived in the community for more than ten years were more likely to report good practices than those who had resided for less than two years (aOR = 2.18, 95% CI: 0.69–6.85, p = 0.041). Other factors, including gender, education level, marital status, occupation, and religion, were not significantly associated with practice level (p>0.05).

**Table 7.** Practices regarding onchocerciasis control (N=250).

| Variable | Category | N | Good practice<br>n (%) | Unadjusted<br>p-value | OR | 95% CI | Adjusted<br>p-value |
| --- | --- | --- | --- | --- | --- | --- | --- |
| Gender | Male | 115 | 103 (89.6) | 0.644 | NA |  |  |
|  | Female | 135 | 123 (91.1) |  |  |  |  |
| Age group | <30 | 60 | 52 (86.7) | 0.032 | 1 |  |  |
|  | 31–50 | 126 | 113 (89.7) |  | 1.32 | 0.51–3.41 | 0.572 |
|  | >50 | 64 | 61 (95.3) |  | 3.13 | 1.04–9.39 | 0.041 |
| Education level | None | 19 | 17 (89.5) | 0.217 | NA |  |  |
|  | Primary | 71 | 64 (90.1) |  |  |  |  |
|  | Secondary | 87 | 79 (90.8) |  |  |  |  |
|  | Tertiary | 73 | 66 (90.4) |  |  |  |  |
| Marital status | Single | 87 | 78 (89.7) | 0.314 | NA |  |  |
|  | Married | 163 | 148 (90.8) |  |  |  |  |
| Occupation | Student | 43 | 38 (88.4) | 0.229 | NA |  |  |
|  | Employed | 46 | 41 (89.1) |  |  |  |  |
|  | Farmer | 124 | 112 (90.3) |  |  |  |  |
|  | Business | 37 | 35 (94.6) |  |  |  |  |
| Religion | Christian | 220 | 198 (90.0) | 0.154 | NA |  |  |
|  | Muslim | 18 | 17 (94.4) |  |  |  |  |
|  | Atheist/Other | 12 | 11 (91.7) |  |  |  |  |
| Endemicity | Hypo | 84 | 80 (95.2) | 0.027 | 1 |  |  |
|  | Meso | 116 | 104 (89.7) |  | 0.45 | 0.14–1.43 | 0.178 |
|  | Hyper | 50 | 42 (84.0) |  | 0.26 | 0.07–0.91 | 0.034 |
| Duration lived in the area | <2 years | 25 | 21 (84.0) | 0.043 | 1 |  |  |
|  | 3–10 years | 51 | 45 (88.2) |  | 1.43 | 0.38–5.35 | 0.596 |
|  | >10 years | 174 | 160 (92.0) |  | 2.18 | 0.69–6.85 | 0.041 |

OR=odds ratio; CI=confidence interval; 1=reference group; \*Significant p values, NA=not applicable

### Onchocerciasis Symptoms and Non-Communicable Diseases

Symptoms of onchocerciasis were reported by 188 respondents (75.2%), while 62 (24.8%) indicated no symptoms (Table 8). The most frequently reported symptoms were itching of the skin or eyes (50.4%), followed by nodules (17.2%), blurred vision (16.8%), and skin changes (2.0%). Symptoms were experienced most often in the morning (25.2%) and afternoon (24.4%), with slightly fewer respondents reporting them at night (22.0%). With regard to non-communicable diseases (NCDs), 81 respondents (32.4%) reported having been diagnosed, while 169 (67.6%) reported no diagnosis. Among those diagnosed, arthritis was the most common (61.7%), followed by hypertension (21.0%), stroke (7.4%), epilepsy (4.9%), and diabetes (4.9%). Family history of NCDs was also common, with 150 respondents (60.0%) indicating at least one case in their families. The most frequently reported conditions were arthritis (18.8%), hypertension (14.0%), diabetes (11.2%), epilepsy (11.2%), and stroke (4.8%).

**Table 8.** Onchocerciasis Symptoms and Non-Communicable Diseases.

| Indicative questions | Response categories | N | n (%) |
| --- | --- | --- | --- |
| Do you have or sometimes present with symptoms of Onchocerciasis? | Yes | 250 | 188(75.2) |
|  | No |  | 62(24.8) |
| Which of the symptoms do you experience? | itching skin/eye | 188 | 126(50.4) |
|  | Nodules |  | 43(17.2) |
|  | Blurred vision |  | 42(16.8) |
|  | Skin changes |  | 5(2.0) |
| What time of the day do you mostly experience symptoms? | Morning | 188 | 66(25.2) |
|  | Afternoon |  | 62(24.4) |
|  | Night |  | 60(22.0) |
| Have you ever been diagnosed with any non-communicable disease? | Yes | 250 | 81(32.4) |
|  | No |  | 169(67.6) |
| If yes, which non-communicable diseases have you been diagnosed with? | Stroke | 81 | 6(7.4) |
|  | Diabetes |  | 4(4.9) |
|  | Hypertension |  | 17(21.0) |
|  | Arthritis |  | 50(61.7) |
|  | Epilepsy |  | 4(4.9) |
| Do you have a family history of any non-communicable disease? | Yes | 250 | 150(60.0) |
|  | No |  | 100(40.0) |
| If yes, which non-communicable diseases have been recorded in your family? | Stroke | 150 | 12(4.8) |
|  | Diabetes |  | 28(11.2) |
|  | Hypertension |  | 35(14.0) |
|  | Arthritis |  | 47(18.8) |
|  | Epilepsy |  | 28(11.2) |

It was observed that, of the 188 respondents with onchocerciasis symptoms, 46 (24.5%) reported having an NCD diagnosis, while 84 (44.7%) indicated a family history of NCDs (Fig 3).

**Fig 3.**
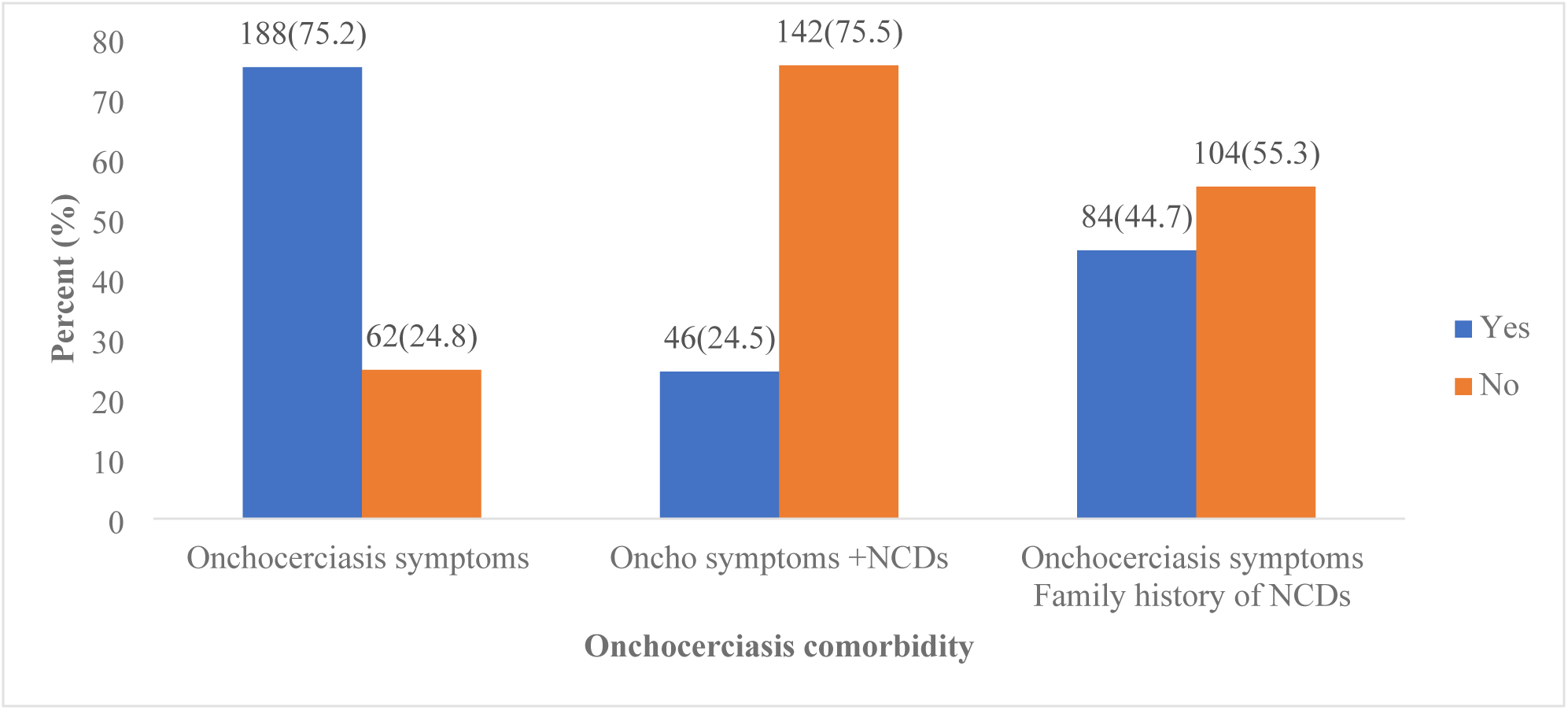
Distribution of onchocerciasis symptoms, NCDs, and family history of NCDs

## Discussion

Despite decades of community-directed treatment with ivermectin (CDTI), onchocerciasis elimination in Cameroon is still being threatened on an unprecedented scale by persistent transmission in endemic foci, suboptimal treatment coverage, the emergence of ivermectin resistance, co-endemicity with other filarial infections, and the additional challenge of rising incidence of non-communicable diseases (NCDs). Onchocerciasis remains endemic across all ten regions of Cameroon, affecting millions of people. Documented barriers include gaps in knowledge, misconceptions about transmission, and weaknesses in the health system, while civil unrest further hinders program implementation [26,27]. Moreover, comorbid conditions such as epilepsy, hypertension, and arthritis increase the clinical burden in endemic areas, complicating case management and elimination efforts. [17,28]. At the same time, the growing prevalence of NCDs, including hypertension, arthritis, diabetes, and epilepsy, places an added strain on communities already burdened by neglected tropical diseases [29]. This study aimed to assess community knowledge, attitudes, and practices toward onchocerciasis and CDTI, document the occurrence of NCDs in the Bafut Health District, and provide an evidence-based guide to integrated approaches for disease control and elimination.

The study shows that while almost all respondents (96.8%) were aware of onchocerciasis, fewer than half (46.0%) correctly identified the filarial worm as the cause. Many attributed the disease to blackflies, mosquitoes, or poor hygiene. Similar misconceptions have been documented in other endemic regions, where communities often confuse the role of blackflies as vectors with that of the parasite itself [10]. Such misunderstandings may reduce confidence in elimination programs, particularly when coupled with the belief in person-to-person transmission, reported by 38.3% of participants, or with fears of ivermectin side effects. These concerns have also been observed in long-term CDTI settings in Cameroon and have been linked to reduced treatment adherence [23]. Targeted community education that clarifies the biological cause of the disease, combined with consistent communication on ivermectin safety, is essential for maintaining adherence and ensuring progress toward elimination.

Our results further reveal a disparity between knowledge and socio-demographic variables. Older respondents and those with higher education levels were significantly more likely to demonstrate a good understanding of onchocerciasis and CDTI. Similar patterns were observed by Kamga *et al*. [30]. In the West Region of Cameroon, individuals with tertiary education displayed a wider understanding of disease causation and prevention. Furthermore, respondents living in hypo-endemic areas were more knowledgeable than those in hyper-endemic zones, echoing findings that endemicity level can shape awareness and risk perception. [25] These disparities underscore the need for targeted interventions that prioritize younger people, those with limited formal education, and residents of high-transmission zones. Making use of community drug distributors (CDDs), who were the primary source of information in this study, may offer a practical pathway to address these gaps. Even so, their communication strategies must be reinforced with accurate, consistent messaging to counteract persistent misconceptions and promote adherence.

The present study revealed mixed community attitudes toward onchocerciasis and CDTI. Fewer than half of respondents perceived onchocerciasis as a severe disease (39.2%) or considered ivermectin effective in preventing it (43.0%). Such findings are consistent with earlier work in Cameroon [10] and other endemic regions in Uganda [31], where low risk perception and doubts about ivermectin’s effectiveness were identified as barriers to sustained participation in the control program, and hindered high coverage and compliance. Misconceptions were also evident, with nearly one-third of participants (32.4%) believing that traditional medicine could cure onchocerciasis, a perception that was similarly reported in studies from Nigeria and Uganda, where reliance on traditional remedies reduced uptake of ivermectin [27,32]. Despite these concerns in the different studies, the majority of participants expressed comfort with taking ivermectin annually (88.3%), and more than half (56.4%) reported that they would encourage family and friends to participate in CDTI.

This study significantly associates positive attitudes with female gender, tertiary education, Muslim faith, and residence in hyperendemic areas. Similar observations were reported elsewhere, where education and other individual characteristics were found to shape long-term compliance with ivermectin distribution programs, with differences noted across gender and ethnic groups [33] In Cameroon, non-compliance has also been linked to prior experiences with side effects, ethnicity, and the number of years an individual has lived in the community [34]. Beyond individual characteristics, community perceptions of how CDTI programs were organized and the commitment of community drug distributors (CDDs) were shown to be more strongly associated with adherence than disease beliefs themselves. [35]. These findings suggest that improving adherence requires correcting misconceptions and strengthening the CDTI implementation system and procedures.

Over 90% of respondents reported prior ivermectin use, and the majority expressed willingness to retake the drug if recommended. However, a large proportion reported side effects, most often itching, joint pain, swelling, or fever, and fear of side effects was the leading reason for refusal among respondents. In Ghana by Otabil *et al*. [36] also showed that adverse reactions were a key barrier to adherence, while in Cameroon, Senyonjo *et al*. [34] identified previous experiences with side effects as predictors of non-compliance. Dissak-Delon *et al*.[35] found that negative perceptions about drug safety undermined confidence in CDTI. Long-term studies further show that prior negative experiences with ivermectin reduce adherence over time, especially when infection risk is perceived as low or when symptoms are absent [33]. In Ethiopia, Ayalew *et al*. highlighted that consistent ivermectin use was more likely when community participation in drug distribution was high and dosing procedures were transparent. These studies emphasize that sustained uptake requires biomedical reassurance, stronger pharmacovigilance, and responsive community engagement to address fears of side effects.

It was further observed that older residents who had lived longer in their communities were more likely to demonstrate good practices, while respondents from hyperendemic areas reported comparatively lower adherence. Comparable results were reported in Ethiopia, where Yirga *et al*. found that compliance was influenced by age, gender, education, and the degree of community trust in the CDTI process. In Cameroon, Senyonjo *et al*.[34] showed that duration of residence was significantly associated with compliance, while Dissak-Delon *et al*. [35] reported that program organization and community drug distributors (CDDs) commitment were stronger predictors of adherence than individual disease beliefs. Duamor *et al*.[39] also observed that weak program delivery and limited community ownership reduced the impact of CDTI in Southwest Cameroon, despite multiple treatment rounds. More recently, evidence from South Sudan confirmed that enhancing awareness campaigns and shifting from annual to biannual treatment markedly improved therapeutic coverage [18]. These findings therefore underscore that strengthening the capacity and reliability of CDDs, improving community ownership, and adapting treatment schedules where needed are essential to sustain high coverage and move closer to elimination targets.

In our study, 75.20% of respondents reported symptoms consistent with onchocerciasis, most commonly itching of the skin or eyes (50.40%), nodules (17.20%), and blurred vision (16.80%). This pattern aligns with known morbidity profiles of onchocerciasis, where skin manifestations and ocular involvement are among the most frequent clinical expressions [40]. Modelling studies suggest that while acute symptoms, such as severe itching, tend to decline more rapidly with long-term ivermectin distribution, chronic conditions like skin changes and ocular damage persist due to cumulative parasite exposure [13]. The observation that many symptoms occurred in the morning and afternoon is consistent with the biting activity of Simulium blackflies, which are most active during daylight hours along fast-flowing river basins [4].

Non-communicable diseases were also frequently reported, with 32.40% of respondents indicating a diagnosis and 60.00% citing a family history. Arthritis was the most common NCD in this cohort, followed by hypertension, stroke, epilepsy, and diabetes, respectively. The comorbidity of onchocercal symptoms with chronic conditions is noteworthy, with nearly one-quarter (24.50%) of symptomatic individuals also reporting an NCD diagnosis, and almost half (44.70%) citing a family history of NCD. While the direct causal links between onchocerciasis and most NCDs remain unclear, recent evidence points to the neurological consequences of chronic infection, including onchocerciasis-associated epilepsy (OAE), which has been increasingly recognised in endemic regions [41]. Chronic immune activation and inflammation related to filarial infection may also interact with pathways contributing to non-communicable conditions. These findings align with the growing recognition that neglected tropical diseases and NCDs are not isolated burdens but may overlap within the same populations, requiring integrated approaches that address both infection control and chronic disease management [42–44].

Given the comorbidity of onchocerciasis with chronic conditions such as arthritis and hypertension in this study, programmatic integration is needed. Linking CDTI platforms with community-based NCD screening and management could address dual disease burdens, improve treatment adherence, and strengthen primary health care. Such integrated approaches would enhance efficiency, reduce stigma, and advance both NTD elimination and NCD control objectives in Cameroon.

## Conclusion

This study highlights that despite widespread awareness of onchocerciasis in the Bafut Health District, persistent misconceptions about disease causation, transmission, and the safety of ivermectin continue to undermine elimination efforts. While ivermectin uptake was encouraging, Fear of side effects and reliance on traditional medicine remain barriers to full participation. Addressing these requires community-tailored pharmacovigilance and sustained health education to strengthen trust in ivermectin and promote adherence. Age, education, religion, and place of residence strongly influenced knowledge, attitudes, and practices. The coexistence of onchocerciasis with non-communicable diseases such as arthritis and hypertension further complicates disease control, adding to the community’s health burden. Sustained progress toward elimination will require targeted education, stronger pharmacovigilance, increased community engagement, and integrated strategies that address infectious and chronic conditions.

## Limitations

This study has some limitations. The cross-sectional design restricts causal interpretation between knowledge, attitudes, practices, and the occurrence of NCDs. Data were self-reported, which may have introduced recall or social desirability bias, particularly for ivermectin use, side effects, and NCD diagnoses; the latter were not clinically verified and may therefore be misclassified. Finally, the study was conducted in a single health district, which may limit generalizability to other endemic settings

## Data Availability

The dataset used in the present study is available from the corresponding author upon reasonable request.

## Acknowledgments

We are grateful to the study participants in the Bafut Health District for their cooperation and willingness to share their experiences. We thank the community leaders and health workers, especially the community drug distributors, for their support during data collection.

## Authors contributions

**Conceptualization:** Irene U. Ajonina-Ekoti (IUA), Marcelus U. Ajonina (MUA)

**Data Curation:** Promise A. Aghaeze (PAA), Theophilus A. Ekoti (TAE), Carine K. Nfor (CKN)

**Formal Analysis:** Marcelus U. Ajonina (MUA), Irene U. Ajonina-Ekoti (IUA)

**Investigation:** Irene U. Ajonina-Ekoti (IUA), Tiburce Gangue (TG), Beri A. Gariba (BAG), Moses A. Mbanwi (MAM), Adolph A. Fozao (AAF)

**Methodology:** Irene U. Ajonina-Ekoti (IUA), Marcelus U. Ajonina (MUA), Promise A. Aghaeze (PAA), Mbunkah D. Achukwi (MDA)

**Software:** Marcelus U. Ajonina (MUA)

**Supervision:** Irene U. Ajonina-Ekoti (IUA), Mbunkah D. Achukwi (MDA)

**Validation:** Irene U. Ajonina-Ekoti (IUA), Marcelus U. Ajonina (MUA), Mbunkah D. Achukwi (MDA), Ebanga Joan (EJ)

**Visualization:** Irene U. Ajonina-Ekoti (IUA), Marcelus U. Ajonina (MUA), Adolph A. Fozao (AAF), Moses A. Mbanwi (MAM)

**Writing – Original Draft Preparation:** Irene U. Ajonina-Ekoti (IUA), Marcelus U. Ajonina (MUA), Theophilus A. Ekoti (TAE)

**Writing – Review & Editing:** Irene U. Ajonina-Ekoti (IUA), Promise A. Aghaeze (PAA), Ebanga Joan (EJ), Theophilus A. Ekoti (TAE), Tiburce Gangue (TG), Beri A. Gariba (BAG), Moses A. Mbanwi (MAM), Adolph A. Fozao (AAF), Mbunkah D. Achukwi (MDA), Carine K. Nfor (CKN), Marcelus U. Ajonina (MUA)

